# Implementing Precision Medicine for Dilated Cardiomyopathy: Insights from The DCM Consortium

**DOI:** 10.1101/2024.11.22.24317816

**Authors:** Elizabeth Jordan, Hanyu Ni, Patricia Parker, Daniel D. Kinnamon, Anjali Owens, Brian Lowes, Chetan Shenoy, Cindy M. Martin, Daniel P. Judge, Daniel P. Fishbein, Douglas Stoller, Elina Minami, Evan Kransdorf, Frank Smart, Garrie J. Haas, Gordon S. Huggins, Gregory A. Ewald, Jamie Diamond, Jane E. Wilcox, Javier Jimenez, Jessica Wang, Jose Tallaj, Mark H. Drazner, Mark Hofmeyer, Matthew T. Wheeler, Omar Wever Pinzon, Palak Shah, Stephen S. Gottlieb, Stuart Katz, Supriya Shore, W. H. Wilson Tang, Ray E. Hershberger, DCM Precision Medicine study of the DCM Consortium

**Author notes:** **Corresponding Author:** Ray E. Hershberger, MD, The Ohio State University Wexner Medical Center, Biomedical Research Tower Room 304, 460 West 12^th^ Avenue, Columbus, OH 43210 USA.

## Abstract

**Background:** Clinical genetic evaluation of dilated cardiomyopathy (DCM) is implemented variably or not at all. Identifying needs and barriers to genetic evaluations will enable strategies to enhance precision medicine care.

**Methods:** An online survey was conducted in June 2024 among cardiologist investigators of the DCM Consortium from US advanced heart failure/transplant (HF/TX) programs to collect demographics, training, program characteristics, genetic evaluation practices for DCM, and implementation needs. An in-person discussion followed.

**Results:** Twenty-five cardiologists (28% female, 12% Hispanic, 68% White) participated in the survey and 15 in the discussion; genetics training backgrounds varied greatly. Clinical genetic testing for DCM was conducted by all programs with annual uptake ranging from 5%-70% (median 25%). Thirteen respondents (52%) did not use selection criteria for testing whereas others selected patients based on specific clinical and family history data. Eight (32%) ordered testing by themselves, and the remainder had testing managed mostly by a genetic counselor or others with genetic expertise (16/17; 94%). Six themes were distilled from open-ended responses regarding thoughts for the future and included access to genetics services, navigating uncertainty, knowledge needs, cost concerns, family-based care barriers, and institutional infrastructure limitations. Following an in-person discussion, four areas were identified for focused effort: improved reimbursement for genetic services, genetic counselor integration with HF/TX teams, improved provider education resources, and more research to find missing heritability and to resolve uncertain results.

**Conclusions:** HF/TX programs have implementation challenges in the provision of DCM genetic evaluations; targeted plans to facilitate precision medicine for DCM are needed.

## INTRODUCTION

Dilated cardiomyopathy (DCM), a leading cause of heart failure (HF) and cardiac transplantation in the United States, has an estimated prevalence of 1:250 individuals.^1,2^ Considerable progress has been made to understand the genetic basis of DCM, and from this a consensus has emerged that clinical genetics should be considered as a core component of the evaluation of DCM patients (probands) and their family members.^3–5^ The clinical and genetic evaluation process for DCM, a practice of precision medicine for DCM, presents abundant opportunity to elevate the quality of care.^6–9^ A comprehensive expert clinical and genetic evaluation is the first step to the implementation of precision medicine for the prevention, management, and consideration of emerging treatments for DCM, including gene- and phenotype-directed therapies.^10–15^

Despite the promise of precision medicine for DCM, the real-world delivery of genetics services for DCM remains highly variable, and in most cases is not done. While commercial laboratories offer cardiomyopathy genetic testing panels, precision medicine care exceeds performing only a genetic test, requiring a broader guideline-recommended^3–5^ approach termed a *genetic evaluation*: the combination of rigorous phenotyping, genetic counseling, genetic testing, expert genetic and cardiovascular interpretation with patient-specific educational and psychological support for the family.^4,16–19^ While optimal genetic evaluation models have been proposed^4,19–21^ the implementation of recommended precision care in DCM has not been rigorously evaluated.

A recent study from the Veredigm database of real world data from over 170 million patients showed that fewer than 1% of DCM patients currently receive genetic testing.^22^ The reasons for this gap in DCM genetic care have been minimally explored. In familial hypercholesterolemia (FH), a Centers of Disease Control Tier 1 condition,^23^ documented barriers to genetic testing have included lack of provider knowledge of guidelines, limited access to counseling services, and billing concerns.^24,25^ Similar themes of knowledge gaps and workforce deficits in addition to operational challenges were also reported in a systematic review inclusive of multiple genetic conditions.^26^ In order to plan initiatives to increase implementation of precision medicine in DCM, an understanding of current care models and practice barriers are needed.

The NHLBI-funded DCM Precision Medicine Study consented 1265 DCM probands and 1781 of their first-degree relatives at clinical sites of the DCM Consortium comprised of leading academic advanced HF/heart transplant (TX) programs across the US.^17^ The study results have emphasized the importance of a family-based genetic evaluation for DCM for both probands and their at-risk family members.^27–30^ In part to seek insight into addressing the above issues, DCM Consortium site principal investigators sought to identify current barriers to performing genetic evaluations for probands with DCM and their families. A needs assessment survey and a stakeholder discussion were conducted to understand current models and barriers to genetic evaluation at the Summer Scientific Symposium of the DCM Consortium in July 2024. Themes from the survey and discussion guided the selection of areas for focused efforts to enhance precision medicine care for DCM.

## METHODS

Current practices and needs required to implement precision medicine in DCM were assessed by an electronic questionnaire completed by co-authors who were current site principal investigators of the DCM Consortium. An in-person discussion followed. Complete methods can be found in Supplemental Material. Access to the data from this assessment can be made upon reasonable request.

## RESULTS

### Characteristics of participants and their institutions

Of 25 participating cardiologists, 28% were female, 12% Hispanic, and 68% White (Table 1). Twenty-four of the 25 cardiologists were HF/TX specialists; one other, a general cardiologist, provided data from their HF/TX program physicians. Clinical cardiovascular training experience varied widely: twelve specified no clinical genetics training, seven had worked ≥ 5 sessions with a GC, three had ≥ 5 sessions with a GC plus a national bootcamp or a ≥1 month genetics rotation during training, one had national bootcamp training only, one had 3 years training experience in a cardiovascular genetics clinic, and one had ≥1 year of clinical genetics training.

**Table 1.** Demographic Characteristics and Training Experiences of DCM Consortium site investigator survey participants.

| Characteristic | Number<br>(N=25) | Percent |
| --- | --- | --- |
| *Demographics* |  |  |
| *Sex*, female | 7 | 28.0 |
| *Ethnicity*, Hispanic | 3 | 12.0 |
| *Race* |  |  |
| Asian | 7 | 28.0 |
| White | 17 | 68.0 |
| Unknown | 1 | 4.0 |
| *Clinical Experiences* |  |  |
| *Specialty* |  |  |
| Advanced heart failure and cardiac transplant | 22 | 88.0 |
| Cardiovascular genetics | 1 | 4.0 |
| General cardiology | 1 | 4.0 |
| Imaging | 1 | 4.0 |
| *Years in practice* |  |  |
| 0-5 | 12 | 48.0 |
| 6-10 | 2 | 8.0 |
| 11-15 | 1 | 4.0 |
| 16-20 | 3 | 12.0 |
| >20 | 7 | 28.0 |

The majority (76%) of participant’s institutions (Figure 1) had ≥10 HF/TX cardiologists; 32% had ≥10 advanced practice providers (APPs). The annual new patient volumes in inpatient and outpatient settings were >250 for 21.7% and 27.3% of institutions, respectively. The DCM probands treated at participating institutions had diverse race and ethnicity backgrounds (Table 2).

**Figure 1.**
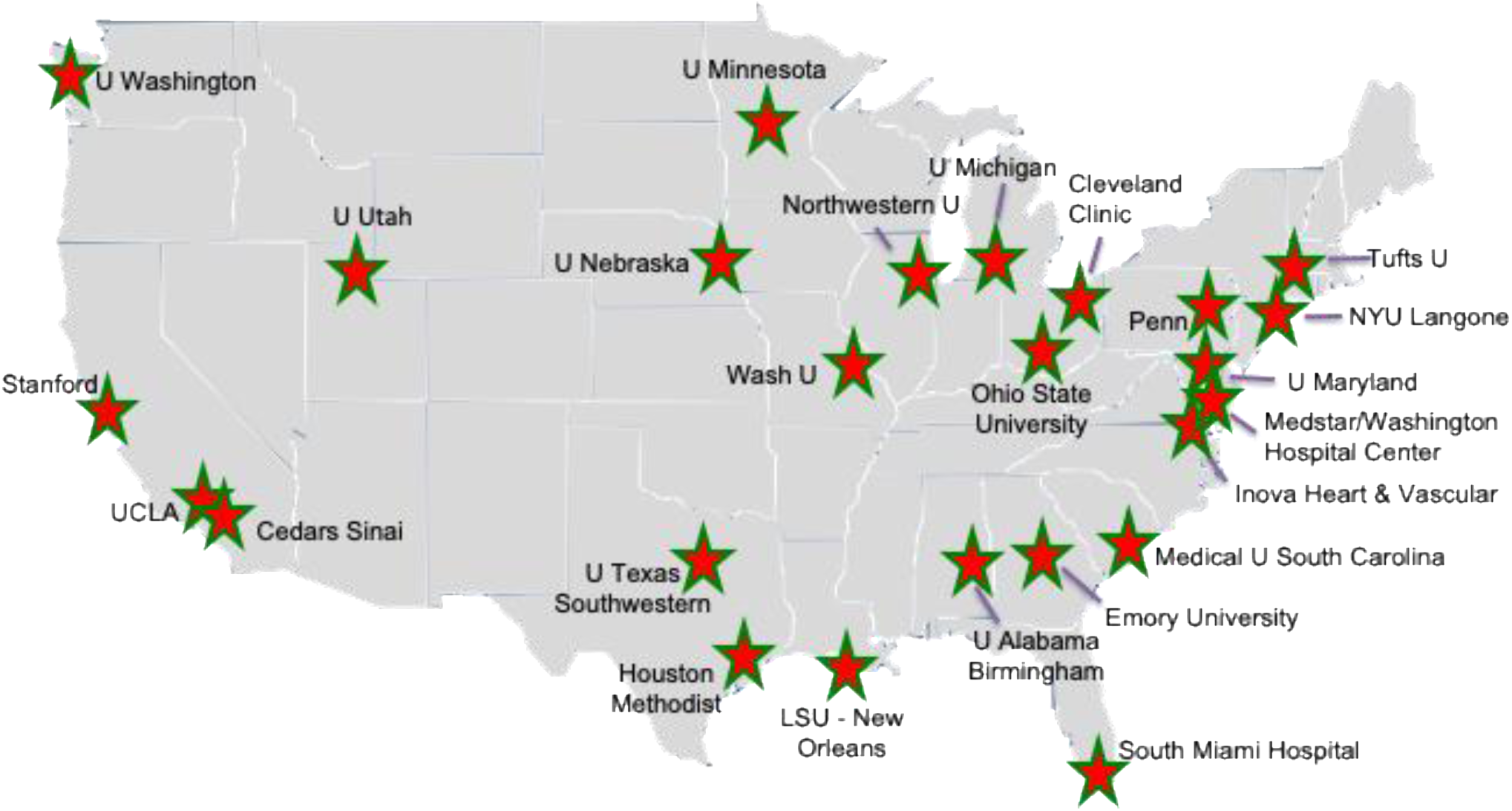
Institutions of the DCM Consortium Site Principal Investigators who participated in the study survey. The map shows the geographic reach of the DCM Consortium.

**Table 2.** Institutional characteristics and annual patient volumes at programs of investigator survey participants.

| Characteristic | Number (N=25) | Percent |
| --- | --- | --- |
| *Number of heart failure/transplant cardiologist and advanced practice providers* |  |  |
| HF/TX cardiologists |  |  |
| <10 | 6 | 24.0 |
| 10-15 | 14 | 56.0 |
| >15 | 5 | 20.0 |
| Advanced practice provider |  |  |
| <10 | 17 | 68.0 |
| 10-15 | 7 | 28.0 |
| >15 | 1 | 4.0 |
| *Annual patient volumes* |  |  |
| New patients* |  |  |
| Inpatient |  |  |
| <100 | 9 | 39.1 |
| 100-250 | 9 | 39.1 |
| >250 | 5 | 21.7 |
| Outpatient |  |  |
| <100 | 2 | 9.1 |
| 100-250 | 14 | 63.6 |
| >250 | 6 | 27.3 |
| Established patients** |  |  |
| Inpatient |  |  |
| <500 | 12 | 54.5 |
| 500-1000 | 6 | 27.3 |
| >1000 | 4 | 18.2 |
| Outpatient |  |  |
| <500 | 2 | 9.1 |
| 500-1000 | 12 | 54.5 |
| >1000 | 8 | 36.4 |
| ***Race/Ethnicity of patients*** |  |  |
| Hispanic*** |  |  |
| <10% | 12 | 52.2 |
| 10-65% | 11 | 47.8 |
| White |  |  |
| <50% | 8 | 32.0 |
| 50-82% | 17 | 68.0 |
| African American |  |  |
| <20% | 10 | 40.0 |
| 20-49% | 11 | 44.0 |
| 50-75% | 4 | 16.0 |
| Asian |  |  |
| <5% | 13 | 52.0 |
| 5-25% | 12 | 48.0 |

\*2 missing values for new inpatients and 3 for new outpatients; \*\*3 missing values for established inpatients and 3 for established outpatients; \*\*\*2 missing values.

### DCM genetic evaluation practices

All indicated that genetic evaluations or testing are conducted at their institutions (Table 3). The estimated annual percentage of DCM probands receiving genetic testing at each program ranged from 5%-70% (average of 26%; Figure 2). Half responded that they did not have selection criteria for DCM patients for genetic evaluation or testing. The remainder based decisions on clinical or family history factors, with the highest ranked clinical criterion as younger age of onset, and highest ranked family history criterion as the presence of multi-generational first- or second-degree relatives with history of DCM or sudden cardiac death (Table 4).

**Figure 2.**
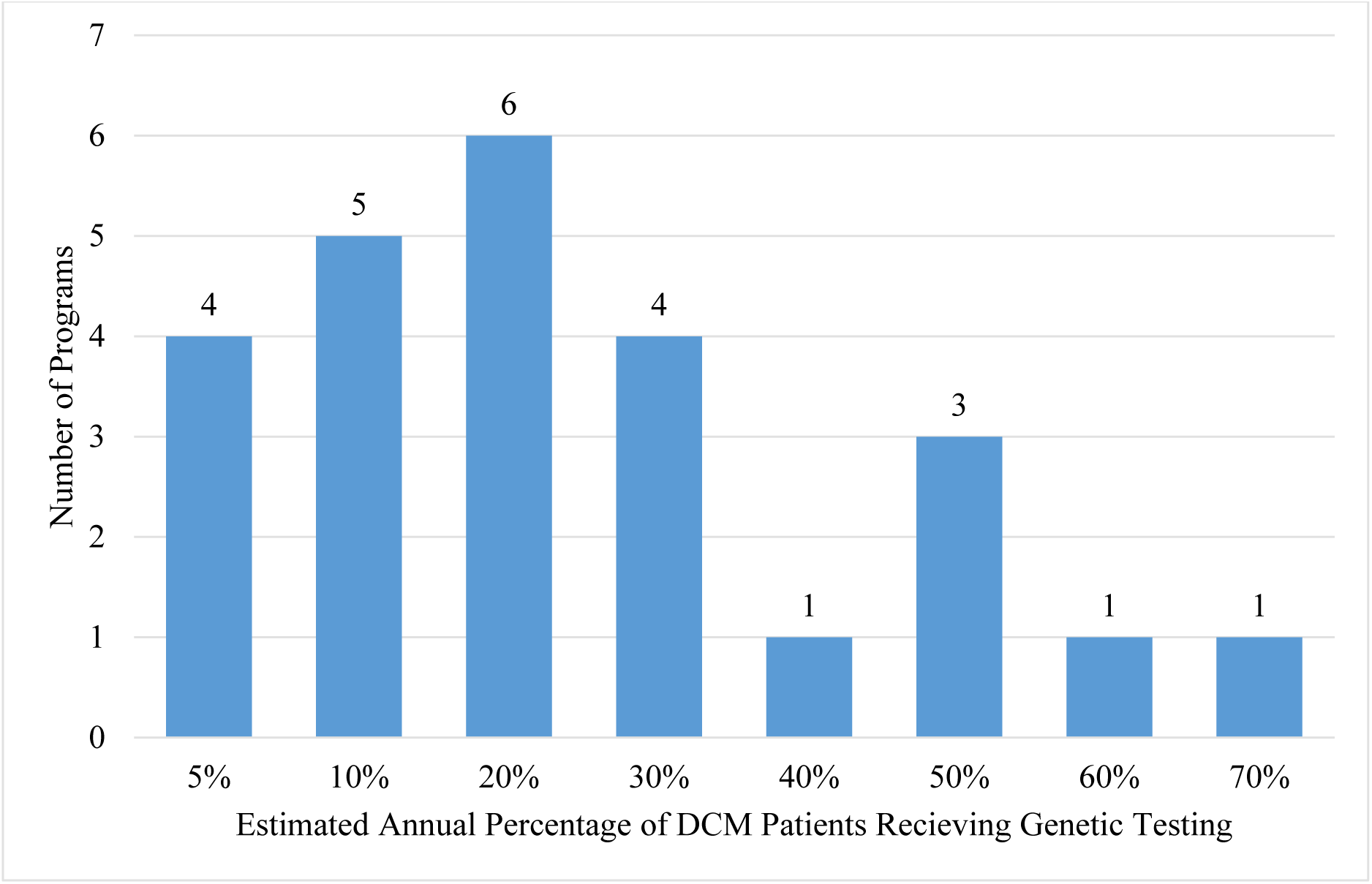
Estimated annual percentage of patients with dilated cardiomyopathy receiving genetic testing at each program. The approximate percentage of DCM patients estimated to receive genetic testing per year at the DCM Consortium sites of the participating investigators as provided in Section B, Question 1 of the survey (Appendix 1).

**Table 3.** Genetic evaluation and testing practices of investigators at sites of The DCM Consortium.

| **Genetic Evaluation/Testing Practice** | **Number**<br>**(N=25)** | **Percent** |
| --- | --- | --- |
| **Are clinical genetic evaluations or genetic testing conducted for DCM patients at your center? (Yes)** | 25 | 100.0 |
| **How do you select DCM patients for genetic testing?** |  |  |
| I test all DCM patients | 13 | 52.0 |
| I test DCM patients based on their clinical or family history data | 12 | 48.0 |
| **Do you conduct DCM genetic evaluation yourself?** |  |  |
| Yes | 8 | 32.0 |
| No, I have others to assist with this | 17 | 68.0 |
| A genetic counselor | 8 | 32.0 |
| A genetic counselor or others with genetics expertise<sup>1</sup> | 8 | 32.0 |
| No one | 1 | 4.0 |
| ***Pre-test Coordination and Genetic Services*** |  |  |
| **Where is the genetic test sent?** |  |  |
| External test vendor | 24 | 96.0 |
| Both internal and external | 1 | 4.0 |
| **Who manages insurance authorizations?** |  |  |
| Administrative or clinical staff | 7 | 28.0 |
| Genetic counselor | 10 | 40.0 |
| MD/self/nurse | 5 | 20.0 |
| Other | 3 | 12.0 |
| **In what clinical setting are you able to obtain genetic testing?** |  |  |
| Inpatients and outpatients | 13 | 52.0 |
| Outpatients only | 12 | 48.0 |
| **Is any pretest genetic counseling provided? (Yes)** | 19 | 76.0 |
| Who does it? |  |  |
| A genetic counselor | 8 | 42.1 |
| You or genetic counselor | 5 | 26.3 |
| Nurse or other or genetic counselor | 6 | 31.6 |
| ***Return of Genetic Test Results*** |  |  |
| **Who at your institution receives genetic results?** |  |  |
| A genetic counselor | 4 | 16.0 |
| You | 4 | 16.0 |
| You and genetic counselor | 8 | 32.0 |
| You or a geneticist or a nurse | 9 | 36.0 |
| **Do you have a process reviewing or evaluating genetic test results?** |  |  |
| Yes | 12 | 48.0 |
| No | 13 | 52.0 |
| **Are genetic results returned to patients? (Yes)** | 25 | 100.0 |
| **How are the results provided?** |  |  |
| In person at clinic | 3 | 12.0 |
| By phone call or a letter or in person | 22 | 88.0 |

| ***Family Follow-up*** |  |  |
| --- | --- | --- |
| **Are implications for family members provided? (Yes)** | 25 | 100.0 |
| **Are first degree relatives offered screening with your program? (Yes)** | 23 | 92.0 |

<sup>1</sup>Others with genetics expertise included cardiologists with genetics expertise, medical geneticists, or an APP or Nurse with genetics expertise.

**Table 4.** Top 5 criteria used by participating investigators to select patients for genetic testing based clinical and family history data.

| **Rank** | **Clinical data criteria** | **Family history criteria** |
| --- | --- | --- |
| 1 | Younger age of onset (24 points) | Multigenerational first- or second-degree relatives with history of DCM or SCD (24 points) |
| 2 | Low LV ejection fraction (12 points) | More than one first degree relative with DCM (20 points) |
| 3 | Larger LV end-diastolic dimension (9 points) | One first-degree relative with established history of DCM (18 points) |
| 4 | Second- or third-degree heart block or presence of a pacemaker (7 points) | One FDR with SCD or sudden unexplained death (10 points) |
| 5 | Rapid progression of DCM and heart failure (5 points) | One first-degree relative with suspicion of DCM (2 points) |

NOTES: Only for those who responded that the selection of patients for genetic testing is based on clinical or family history data (n=12). Ranking is based on total points calculated. Points were determined by the rank orderings of top 3 criteria (3 points for top 1, 2 points for top 2, and 1 point for top 3) multiplied by the frequency at each ranking, respectively. The top five with highest cumulative score for each criteria category are shown.

Eight participants (32%) conducted genetic evaluations themselves. Of those that have other team members to assist (n=17), 94% reported that the genetic evaluation was usually conducted by GCs or others with genetic expertise (Table 3). Genetic testing insurance authorizations were mostly handled by GCs (40%) followed by administrative assistants or clinical staff (28%). Two-thirds of the programs provided pretest counseling, either by GCs, themselves, or a nurse. All institutions returned results to patients, with multiple approaches of in person or phone call or letter. Forty-eight percent had a process to review genetic results, with most common approaches including collaborative review along with GCs, geneticists, and/or cardiologists. Implications for at-risk relatives were provided by all programs and most also offered family screening.

### Challenges in conducting a DCM genetic evaluation

Themes regarding major issues and barriers to conducting a genetic evaluation for DCM were identified in the open-ended responses from Section Three (“Thoughts for the Future”) of the questionnaire (Appendix 1). Six common themes emerged (Table S1). In order from most to least commonly cited, themes included lack of access to genetics services (including counseling and testing; n=11 comments), operational and institutional infrastructure limitations (including variable physician uptake, referral coordination, lack of financial/personnel support; n=10), cost concerns (reimbursement of genetic testing and counseling services; n=9), physician education needs (keeping up with the changing knowledge and educating patients on genetic risk; n=7), challenges of family-based care (encouraging family communication, eliciting family history; n=6), and difficulty managing genetic uncertainty (n=5) (Appendix 2; Table S1).

### Areas to focus efforts for future enhancement of precision medicine care in DCM

Four key focus areas for future efforts targeted to improve implementation of a clinical genetic evaluation in DCM were distilled from the themes from the open-ended questionnaire responses regarding “Thoughts for the Future” and the transcription of the in-person discussion (Supplemental Material).

#### Focus Area 1: Improve reimbursement for genetics services for cardiomyopathy

Reimbursement issues were two-fold, including genetic testing and genetic counseling services, with limited and inconsistent payor coverage for both. This was of considerable concern, with one investigator explaining that “*The biggest barrier to DCM genetics is the capacity to have genetic testing covered by insurance and the ease to figure out coverage issues. Many of the patients are unable to pay out of pocket for testing and also providers have avoided ordering genetic testing due to challenges figuring out coverage feasibility.*”

Only a minority of payors have had coverage policies for genetic testing for the indication of cardiomyopathy. Many laboratories now conduct disease-specific tests on an exome or genome backbone but, rather than billing for the true cost of the test, labs have often implemented billing structures designed to meet payor demands.^31,32^ Intermittent pharma-sponsored testing programs have provided temporary relief to remove cost barriers, however avoidance of addressing the lack of reimbursement mechanisms has contributed to unsustainable billing models and limited growth in the field.^31^ The testing landscape continues to evolve, with first-tier cardiomyopathy genetic testing moving toward clinical genome,^33^ however appropriate utilization will be key to maximizing cost effectiveness.

Genetic counseling services also have been poorly reimbursed, if at all, which has limited hiring in clinical care settings. As shown in a recent downstream revenue analysis in pediatric cardiovascular disease,^34^ GCs have contributed to financial success of their institution through reimbursements from downstream family genetic testing, clinical screening, and subsequent ongoing care needs resulting from a genetic evaluation. Beyond cardiovascular disease including cardiomyopathy,^34,35^ this has also been demonstrated in hereditary cancer evaluations.^36^ To directly address the issue of GC service reimbursement, a clear need to engage in advocacy at the national level for payor recognition for GC services was a strong sentiment raised in the symposium discussion. Cost concerns will remain a barrier to realizing precision medicine care until the importance of genetic services is recognized by payors, including the Centers for Medicare and Medicaid Services (CMS).^37^

#### Focus Area 2: Integrate cardiovascular genetic counselors into the heart failure team

Genetic counselors facilitate genetic testing, help patients understand and adapt to medical, psychological, and familial implications of genetic disease, and coordinate family-based follow up. The American Heart Association endorses GCs as vital members of the cardiovascular care team and supports policies that promote access to GCs for all patients and their families with cardiovascular indications.^38^ For over a decade, GCs have been acknowledged as a key part of the multidisciplinary cardiovascular team^3,4,19,21,38,39^ yet access to genetics services was the most common theme among implementation issues (Table S1). One participant rated limited GC support as a “*major issue”* stating, “*We do not have the bandwidth currently from a [genetic counselor] standpoint to provide counseling and testing for all DCM patients who are being seen in our [heart failure/transplant/ventricular assist device] clinics.”*

Beyond the eight centers where the HF/TX physician performed genetic evaluations themselves, GCs or other genetics professionals were typically the ones who facilitated this service. In addition to counseling and testing, a GC integrated into the HF/TX team can mitigate challenges with others aspects of the evaluation, including navigating insurance issues, providing education and support, and facilitating family follow up.^35,40^ Some participants shared that their institutions had GCs, but they were administratively placed in non-cardiovascular service areas such as obstetrics or oncology and already had full clinical loads, making it impractical to meet the needs of the HF/TX team. Accordingly, a need for a GC specifically dedicated to the cardiovascular specialty was discussed as the optimal arrangement. As shown in a national cardiovascular case series, when a GC was integrated directly into a cardiology practice, improvements were seen in applying the most appropriate testing strategy and in the accuracy of result interpretation for management purposes.^19^

The GC is also an institutional resource for novel genetic service delivery models. This could include adapting the traditional outpatient model to increase throughput with involvement of APPs in addition to developing workflows beyond the outpatient setting,^41^ with substantial DCM inpatient volumes reported at consortium sites. This is particularly relevant to DCM where advanced stage disease has been associated with P/LP genetic backgrounds.^30^ Existing inpatient models at DCM Consortium sites and a need for in-house genetic testing services to facilitate inpatient genetic evaluation were emphasized in participant comments and discussion (Table S1).

Although guidelines and professional societies have recommended genetic counseling as a part of the genetic evaluation for cardiomyopathy, integration of GCs into cardiovascular care has remained suboptimal. In contrast, there has been greater utilization of clinical genetic evaluation for heritable cancer risk, with the National Comprehensive Cancer Network requiring genetic services for several primary tumor types and/or early diagnoses.^42^ Further, to acquire accreditation as a quality oncology care facility, provision of appropriate genetics services must be demonstrated.^43^ Heart failure centers lack similar requirements, potentially limiting the prioritization of resources to routinely implement genetic evaluation for DCM in the HF community.

#### Focus Area 3: Provide DCM genetics and genomics education resources for heart failure providers

A broad range of education needs was reported, from aiming to simply stay current with the evolving genomics landscape to explaining genetic risk information to patients. These explanations were described as particularly vexing for those surrounding genetic uncertainty, with participants summarizing that “*a complex VUS is not something that most cardiologists are comfortable with communicating or handling*” and how this challenge was deepened by “*poor literacy in some patients.*” An explicit request for “*formalizing training/support for cardiac genetics for HF providers*” was suggested in order keep pace with continued progress in DCM clinical genetics.

The reported educational concerns were consistent with a prior survey of 131 cardiology and electrophysiology practitioners that found that more than half of providers were not confident about genetic testing options available or how to order tests.^44^ Topics sought by nearly all of the providers were translational in nature, such as how to perform a genetic risk assessment and understanding guideline-based management recommendations.^44^ The content required to provide this practical information will continue to evolve, particularly as genome sequencing becomes more accessible, introducing the need to more routinely manage secondary and incidental findings, evaluate test quality, and manage evolving billing and reimbursement structures.

Reported physician knowledge gaps were commonly rooted in managing clinical genetic uncertainty. With about half of DCM patients having VUSs reported at testing,^29^ and many genes of uncertain significance having only limited evidence included on cardiomyopathy panels,^45^ clinicians are often faced with inconclusive information for which the implications are unknown. An interview series of 29 cardiologists reflected how uncertainty disrupts the diagnostic process.^46^ Some have argued that smaller, phenotype-targeted panels with only high evidence genes are favorable in order to reduce the interaction with uncertain results. However, a more sustainable approach may be to build a multi-disciplinary team with clinical genetics expertise for both physicians and HF APPs to remain knowledgeable of the continuously changing genomics landscape. With successful integration of GCs into the HF team (Focus Area 2), this will directly facilitate the cited educational requests, as GCs are able to support and provide expertise for not only patients but also partner providers (Figure 3).

**Figure 3.**
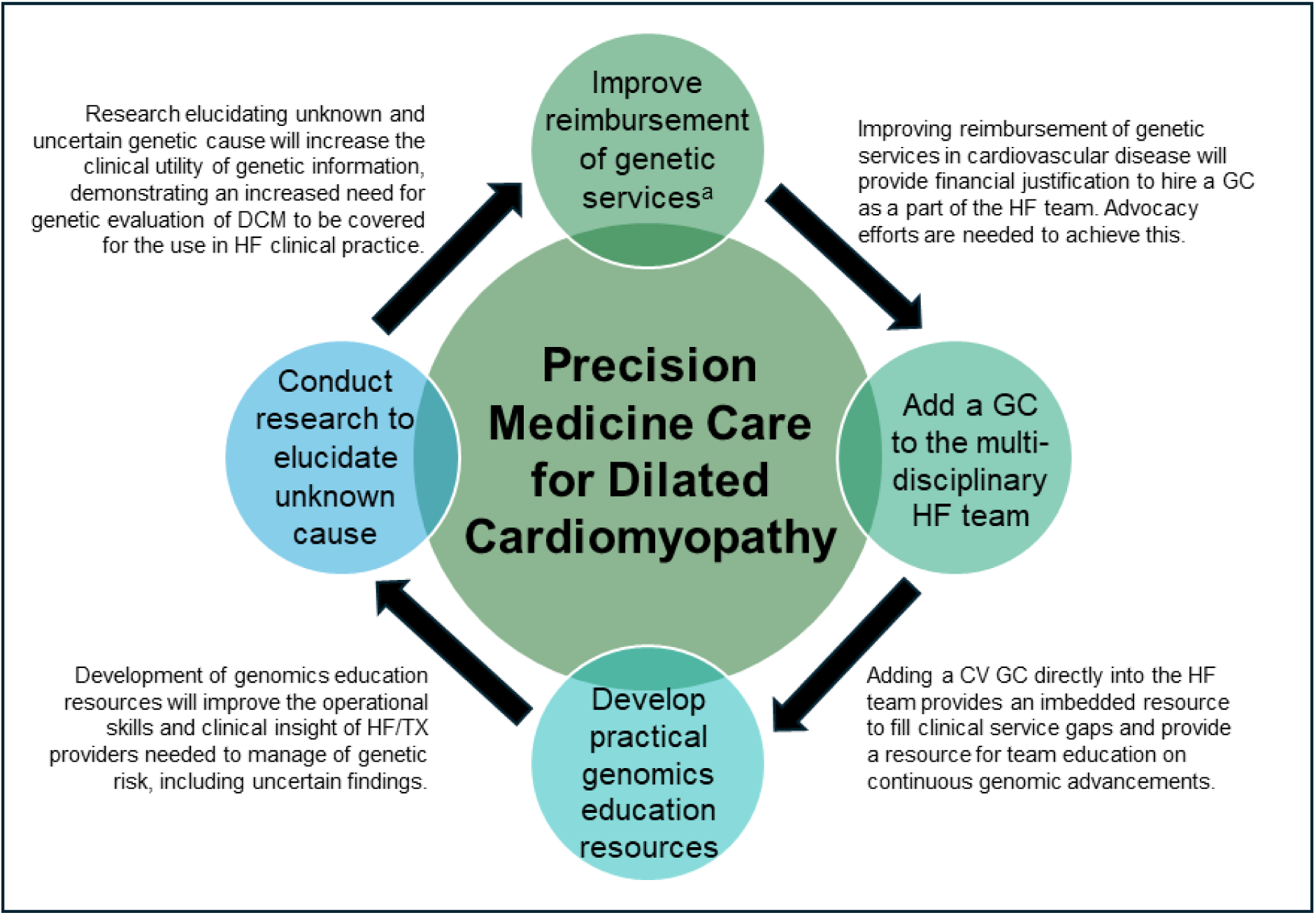
Focused efforts will work synergistically to enhance precision medicine care for dilated cardiomyopathy. This figure summarizes insights of the participating investigators. Resolving reimbursement challenges was considered a top priority. Increased utilization of clinical genetics will enhance implementation opportunities. CV = Cardiovascular; GC = Genetic Counselor; HF = heart failure. ^a^Genetic services include genetic testing and genetic counseling.

#### Focus Area 4: Research to elucidate unknown cause and clarify genetic uncertainty in cardiomyopathy

*“The challenge of handling VUSs is a substantial problem.*” Genetic uncertainty was among the most common themes related to challenges for the future implementation of precision medicine in DCM. This barrier was not misplaced, as substantial genetic complexity for DCM remains.^2,47^ Illustrating this, low rates of actionable P/LP results (estimated 8.2-32.6%) and high rates of VUS (28.3-49.3%) were observed in the Precision Medicine study, with the odds of a P/LP variant in probands with at least one rare variant observed at an estimated 75% lower odds among those of African genomic ancestry compared with those of European ancestry.^29^ Resolution of this discrepancy in identification of actionable genetic results will be required to improve opportunities for care for all. Until resolved, the clinical management of uncertain genetic findings will continue to be a barrier to future implementation of gene specific therapy where well-informed assessment of variant pathogenicity and defining molecular mechanisms will be critical.

As one participant stated, there is a “*need [for] more research to determine variants…or genes that contribute to missing heritability of DCM.*” Additional unidentified variants in non-coding regions or undiscovered genes may be at play, as well as mechanisms that exceed the monogenic paradigm, including oligogenic and polygenic effects and/or contribution from other modifying factors.^47^ Addressing the uncertainty problem is not only an issue of clinical and scientific relevance, but also an issue of equity, as the current picture of DCM genetics has largely been derived from studies of European ancestry, contributing to a currently incomplete picture of DCM genetic architecture.^29^ Large-scale, family-based studies of diverse populations will be required to elucidate currently unsolved cause for all.

## DISCUSSION

This report has provided a contemporary assessment of current precision medicine practice for DCM by cardiologists at advanced HF/TX programs at DCM Consortium sites across the United States. The participating investigators estimated that on average about a quarter of DCM patients were offered genetic testing at DCM Consortium sites, a much higher rate than the <1% uptake estimated for US providers for all of heritable cardiovascular disesase.^22^ This higher rate of clinical genetics practice provided experience and insight to identify barriers and opportunities for DCM precision medicine.

While the full potential of precision medicine care for DCM has yet to be realized, DCM Consortium investigators recognized that integrating clinical genetics into DCM care will enhance precision medicine opportunities, with one participant aspiring that “*Genetic testing should become mainstream as the expertise in the area grows and new therapies become available.”* Accordingly, the collective insight revealed in this assessment may be helpful to plan efforts to more fully integrate genetic evaluations into HF/TX practice.

The multiple challenges summarized by investigators will require broadly-based actions to address financial, personnel, educational, and investigative needs, as described in the four Focus Areas distilled from this needs assessment. Strategic efforts focused in the proposed areas can work synergistically to enhance precision medicine care for DCM (Figure 3). However, other stakeholders beyond HF/TX cardiologists will need to be engaged, including laboratory personnel, payors, legislators, healthcare administrators, and genetic counselors. With collaboration of these key groups, current barriers to implementing precision medicine for DCM are surmountable and, assuming success, will enable hope for a future of improved therapeutic and prevention strategies for HF in DCM probands and their families.

## STRENGTHS AND LIMITATIONS

Strengths of this assessment include the multi-center involvement of geographically dispersed HF programs and the combined quantitative and qualitative questions that provided authentic perspectives on current practices from key physician stakeholders. Limitations include the small sample size from selected advanced HF/TX clinics and that data regarding each program was represented by one physician from each institution, affecting generalizability. In addition, the data were self-reported by individual providers engaged in the field of DCM genetics as investigators in the DCM Consortium, which may represent more advanced genetic evaluation practices relative to others. However, the experience of the participating cardiologists provides direct insight to identify barriers to implementing clinical genetics and precision medicine care, which can be instructive for the broader HF community.

## Data Availability

Data from this manuscript can be shared upon reasonable request.

